# The Japanese version of the Musculoskeletal Pain Intensity and Interference Questionnaire for Musicians (MPIIQM-J): Translation, cultural adaptation, and validation in Higher Music Education Institutions

**DOI:** 10.64898/2026.08.18.26360635

**Authors:** Miki Akaike, Tomoko Tanaka, Yoshimi Suzukamo

**Affiliations:** University of Yamanashi; Senzoku Gakuen College of Music; Ritsumeikan University

**Keywords:** musicians, play-related musculoskeletal disorders, cross-cultural adaptation, questionnaire

## Abstract

**Background:** Playing-related musculoskeletal disorders are highly prevalent among musicians, yet no validated The aim of this study was to translate and culturally adapt the MPIIQM into Japanese and to examine its psychometric properties in students of higher music education institutions (HMEIs).

**Methods:** Forward–backward translation followed ISPOR guidelines, with linguistic validation through pilot testing. Psychometric evaluation was conducted in instrumental music majors enrolled at nine HMEIs in Japan, combining in-person and online surveys. Exploratory factor analysis, Cronbach’s α, and Pearson correlations with the Brief Pain Inventory (BPI), QuickDASH, and QuickDASH Performing Arts Module (PAM) were computed in symptomatic respondents (n = 48). Symptom prevalence in the full HMEIs sample (n = 226) was compared with national data (Ministry of Health, Labour and Welfare in Japan, 2022) using binomial tests with Holm correction.

**Results:** Exploratory factor analysis yielded a two-factor structure (pain intensity; pain interference) explaining 76.8% of the variance. Cronbach’s α was .864 (pain intensity), .890 (pain interference), and .903 (total). The pain intensity subscale correlated strongly with BPI pain severity (r = .71), and the pain interference subscale correlated with the QuickDASH PAM (r = .62). Test–retest reliability could not be computed because only 9 of 30 retest respondents met inclusion criteria. HMEIs students reported significantly higher prevalence than national peers in 25 of 41 symptoms, most markedly stiff shoulders and low back pain.

**Conclusions:** The MPIIQM-J demonstrated adequate structural validity, internal consistency, and convergent validity, and is expected to be useful in educational and clinical settings.

## 2. Introduction

Playing-related musculoskeletal disorders (PRMDs) have been reported at high prevalence rates among musicians (Steinmetz, 2016). Although interventions such as strength training and education for prevention have demonstrated short-term benefits, long-term improvement strategies remain a challenge (Laseur et al., 2023).

In Japan, Enami (1892) was the first to report the existence of occupational palsy among musicians. Subsequently, Seo (1901) reported the treatment of eight koto players (a traditional Japanese instrument) presenting with paralytic symptoms using massage therapy. Following the report by Sakai et al. (1991) of 37 cases of upper extremity pain among pianists, the accumulation of epidemiological research has progressed steadily; since 2000 alone, 21 population-based epidemiological studies have been published. A review by Akaike (2026) of these studies found that 14 of the 21 studies (66.7%) reported prevalence rates of 50% or higher, with 5 of those 14 (35.7%) reporting particularly high prevalence rates of approximately 80%. Specifically, Saito et al. (2006) reported a prevalence of 85% among professional musicians (79% in males and 92% in females), while Kawasaki et al. (2007) found that 85.8% of 267 music university students reported pain. Furuya et al. (2006), in a study of Japanese female pianists ranging from high school students to university students and professional pianists, reported a prevalence of 77%, with more than four hours of daily practice, a neurotic personality, and dedication to practice suggested as risk factors. Kanazuka et al. (2015) also reported a high prevalence of 84%, with the upper extremities (49%), lower back (19%), and cervical region (18%) identified as the primary sites of symptom occurrence. In contrast, Tanaka et al. (2018), conducted among undergraduate and graduate students at Tokyo University of the Arts—an institution with particularly high-performance demands—reported a notably lower prevalence of 13.4–15.3%, highlighting that the appropriateness of questionnaire items and the level of performers’ technical proficiency may influence prevalence estimates.

Although these studies collectively demonstrate that PRMDs represent a significant occupational health concern among musicians in Japan, the majority of them cite insufficient evidence or limited understanding of the actual situation as their research rationale, suggesting that the development of validated musician-specific patient-reported outcome measures (PROMs) is urgently needed.

Against this background, the MPIIQM was developed in English by Berque et al. (2014) to address the dual challenge of unvalidated psychometric properties in self-report measures widely used in PRMD epidemiological research and the absence of a validated musician-specific outcome measure. The development process employed rigorous methodology, including a comprehensive literature review of musician-specific PROMs, adherence to the World Health Organization (WHO) International Classification of Functioning, Disability and Health (ICF) guidelines, the convening of an expert panel for content validity, and evaluation of comprehensive psychometric properties. The questionnaire comprises 22 items encompassing demographic information (2 items), music-related factors (6 items), PRMD prevalence (4 items), a body chart indicating pain location, and two subscales assessing pain intensity and pain interference.

The MPIIQM has since undergone cultural adaptation and validation into seven languages: German (Möller et al., 2018), Polish (Cygańska et al., 2021), Brazilian Portuguese (Kochem & Silva, 2021), French (Roos et al., 2023), European Portuguese (Zão et al., 2023), Italian (Panuccio et al., 2024), and Chinese (Fu et al., 2026).

Across these adaptations, favorable psychometric properties have been consistently reported, including support for content validity and structural validity, and high internal consistency (Cronbach’s α = 0.78–0.92). The MPIIQM has thus become established as the standard musician-specific PROM in international research, education, and clinical settings.

In Japan, where the estimated total musician population is approximately 1.14 million, no Japanese-language version of the MPIIQM is currently available (Akaike,2026). This represents a significant barrier for Japanese-speaking clinicians, educators, and researchers seeking to utilize the MPIIQM in clinical practice, education, and research.

The aim of the present study was to translate and culturally adapt the MPIIQM into Japanese and to examine its validity and reliability as a measurement scale.

## 3. Methods

### 2.1. Ethics

This study was conducted with permission for the development of the Japanese version granted by the original author, Patrice Berque, in April 2023. The study was also conducted in compliance with the Declaration of Helsinki and the Ethical Guidelines for Medical and Health Research Involving Human Subjects, following approval by the Ethics Committee of the Faculty of Medicine, University of Yamanashi (Approval No. 2851). Written informed consent was obtained from all participants prior to their enrollment in the study.

### 2.2. Translation and Cultural Adaptation

Translation and cultural adaptation were conducted in accordance with the guidelines for translation and cultural adaptation of patient-reported outcome measures outlined by Wild et al. (2005). The entire translation process was carried out by four specialists. All translators were native speakers of Japanese; two were researchers with a background as performing musicians, one was a specialist in Japanese-language scale development, and one was a professional translator.

### 2.3. Modification of the Body Chart

The body chart included in the MPIIQM-J was adapted from the version used by Zão et al. (2023), as the original MPIIQM did not include a detailed body diagram. While some online survey platforms offer an interactive body chart that allows respondents to directly select anatomical regions on the screen, such functionality is not yet widely implemented or consistently available across survey environments. Therefore, in order to ensure compatibility with both paper-based and online survey formats, a body chart with clearly numbered anatomical regions was adopted. The selected body chart provides numbered anatomical regions and covers 92 distinct body areas, allowing for more precise and standardized identification of symptom locations regardless of the mode of administration. The use of this modified body chart was approved in advance by the original author of the MPIIQM.

### 2.4. Forward Translation

Two researchers independently translated the original English version of the MPIIQM into Japanese. The two translations were then reviewed by a panel of specialists to assess semantic, idiomatic, experiential, and conceptual equivalence, and a reconciled version was produced.

### 2.5. Back Translation

One professional translator who was blinded to the original version performed a back-translation of the reconciled version into English. The back-translation was then reviewed and finalized by the remaining three specialists. The original author subsequently reviewed the resulting back-translation.

### 2.6. Pilot Testing

To confirm linguistic validity, a pilot test and debriefing interviews were conducted with professional instrumental musicians whose native language was Japanese. Participants were first asked to complete the questionnaire, after which verbal feedback was collected regarding the degree of comprehension and the appropriateness of expression for each item in Japanese. The feedback was reviewed by three specialists, and revisions were made where necessary before the final Japanese version (MPIIQM-J) was established.

### 2.7 Participants

The survey was administered in two separate rounds. Participants in the first round were native speakers of Japanese enrolled in undergraduate or graduate programs at higher music education institutions (HMEIs) in Japan, majoring in instrumental music. Written informed consent to participate in the study was required as a condition of participation. Participants in the second round met the same eligibility criteria as the first round, with the additional requirement that they were currently experiencing or had experienced performance-related symptoms within the past month. According to Zão et al. (2023), the MPIIQM was designed and validated for use with professional orchestral musicians, and its validity has not been established for non-professional or non-orchestral musicians. However, upon consulting the original author regarding the intended scope of application for the present study, it was confirmed that the instrument is not necessarily restricted to professional orchestral musicians and may be applied to other musician populations, including amateur musicians and students in HMEIs. Furthermore, students were included among the respondents in the MPIIQM-F (Roos et al., 2023). Therefore, while it should be noted that existing validation studies have been conducted primarily with professional orchestral musicians, the MPIIQM is considered to have applicability to a broader range of musician populations.

### 2.8. Data Collection

Data collection was also conducted in two separate rounds. The first round was carried out between September and November 2025 at four higher music education institutions in Japan. Booths for questionnaire completion were set up on each HMEIs campus, and participation was voluntary; however, participants who completed the questionnaire received a 500-yen gift card. As the first round did not yield a sufficient sample size for factor analysis, a supplementary online survey was conducted between January and March 2026 using Google Forms. Participants in the online survey likewise received a 500-yen gift card upon completion. To assess test-retest reliability, participants from the first round who volunteered were asked to complete the same questionnaire again three to five days after their initial response. In addition to the MPIIQM-J, each participant simultaneously completed the Japanese version of the Brief Pain Inventory (BPI), the Japanese version of the QuickDASH, and the QuickDASH Performing Arts Module as comparator measures for the assessment of convergent validity. Furthermore, to compare participants’ health status with the national average in Japan, items were drawn from the Ministry of Health, Labour and Welfare’s Comprehensive Survey of Living Conditions, specifically the symptom prevalence question in which respondents select from 41 items any subjective symptoms experienced over the preceding few days (multiple responses permitted).

All responses were collected independently, and participants were not exposed to other participants’ responses. For test–retest assessment, responses were obtained without access to prior answers.

### 2.9. Statistical Analysis

All analyses were conducted using Python (version 3.12.3), with a significance level of 5%. The following Python libraries were used: factor_analyzer (version 0.5.1) for exploratory factor analysis, and SciPy (version 1.17.1) for power analysis, the Mann-Whitney U test, Pearson correlation coefficients, and binomial tests.

Only participants who answered “yes” to MPIIQM items Q11 or Q12 were included in the analysis.

#### ① Sample Size

The target sample size was set at 63 based on the COSMIN checklist. A post-hoc power analysis was also conducted using Python (SciPy version 1.17.1). The effect size (f^2^ = .390) was calculated from the mean inter-item correlation coefficient among the nine MPIIQM items (r = .530), and statistical power was estimated based on the F-distribution at a significance level of α = .05.

#### ② Missing Data

The proportion of missing values for all items of the MPIIQM-J was calculated and evaluated by comparison against the acceptable threshold reported in the original publication (less than 3%).

#### ③ Structural Validity

Prior to conducting factor analysis, the suitability of the data for factor analysis was confirmed using Bartlett’s test of sphericity and the Kaiser-Meyer-Olkin (KMO) measure of sampling adequacy. To assess structural validity, exploratory factor analysis (EFA) was conducted using the principal axis factoring (PAF) method. The number of factors was determined using the Kaiser criterion (eigenvalue ≥ 1), and Varimax rotation was applied. A factor loading of 0.40 or above was adopted as the criterion for significance.

#### ④ Internal Consistency

Internal consistency was calculated using Cronbach’s α for each subscale and the overall scale. An α value between 0.70 and 0.90 was adopted as the criterion for acceptable internal consistency. Additionally, corrected item-total correlations and changes in α upon deletion of each item were examined.

#### ⑤ Test-Retest Reliability

The intraclass correlation coefficient (ICC) was calculated using the data obtained in the first round and the 30 paired retest datasets subsequently collected.

#### ⑥ Convergent Validity

To assess convergent validity, Pearson product-moment correlation coefficients were calculated between each subscale of the MPIIQM-J and the BPI, QuickDASH, and QuickDASH Performing Arts Module. A priori hypotheses were that a strong correlation (r > 0.70) would be observed between the MPIIQM-J pain intensity subscale and BPI pain intensity, and that a moderate to strong correlation (r = 0.50–0.70) would be observed between the MPIIQM-J pain interference subscale and the QuickDASH Performing Arts Module.

#### ⑦ Comparison between In-person and Online Survey

To examine differences in item scores between the two data collection methods (in-person vs. online), the Mann-Whitney U test was conducted, with Holm correction applied for multiple comparisons. Effect size r was calculated.

#### ⑧ Comparison among Instrument Groups

To exploratorily examine differences among instrument groups, Kruskal–Wallis tests were conducted for age, BMI, MPIIQM-J pain intensity, pain interference, total score, and the number of symptoms reported in the Comprehensive Survey of Living Conditions.

#### ⑨ Descriptive Analysis

For symptom prevalence, a descriptive comparison was made with the published values for the 20–24 age group from the Comprehensive Survey of Living Conditions. Binomial tests were conducted to compare the symptom prevalence observed among students in the survey against the national population proportions for each symptom as the reference, with Holm correction applied for multiple comparisons.

## 3. Results

### 3.1. Translation and Cultural Adaptation

In both the forward translation and back-translation, discrepancies between translations were minor and resolved by consensus among the panel members. The pilot test conducted in November 2023 involved five professional instrumental musicians (mean age 33.8 years, SD = 6.69; three females, two males). The instruments played by participants were trumpet, flute, cello, piano, and Tsugaru-jamisen, representing a diverse range that encompassed both Western and traditional Japanese instruments. The purpose of the pilot test was to confirm linguistic validity, and the eligibility criterion was being a native Japanese-speaking instrumental musician, regardless of instrument type. The mean completion time for the questionnaire was 8 minutes 19 seconds (SD = 2 minutes 38 seconds). Following a review of the feedback collected on the comprehensibility and appropriateness of expression of each item in Japanese, the MPIIQM-J was finalized after minor wording revisions.

### 3.2. Participant

In the first round of in-person data collection (September–November 2025), questionnaires were administered at four higher music education institutions in Japan, yielding responses from 200 participants. Of these, two participants whose primary major was not instrumental music were excluded. Among the remaining participants, 27 responded “yes” to either Q11 (presence of pain interfering with playing during the last month) or Q12 (presence of similar pain within the past seven days) of the MPIIQM. A second round of online data collection was therefore conducted (January–March 2026), yielding an additional 26 responses, bringing the total to 53 participants. Of these, five were excluded due to response inconsistencies, resulting in a final analytical dataset of 48 participants. The characteristics of the 48 participants included in the analysis were as follows. Academic level ranged from first-year undergraduate to first-year doctoral students, with a mean age of 21.31 years (SD = 1.74). Gender distribution was 37 females, 9 males, and 2 unreported. Mean BMI was 20.41 (SD = 2.05). Participants were enrolled at nine institutions: five private HMEIs (a: n = 10, b: n = 14, c: n = 3, d: n = 1, e: n = 1) and four national HMEIs (f: n = 5, g: n = 9, h: n = 4, i: n = 1).

### 3.3 Statistical Analysis

#### ① Sample Size Adequacy

The statistical power for the obtained sample size (n = 48) was .812, exceeding the conventionally accepted threshold of 1 − β = .80. The minimum sample sizes required to achieve power levels of .80, .90, and .95 were 47, 58, and 67, respectively, indicating that the present sample met the criterion for power of .80. Based on these results, the sample size of the present study was deemed adequate in terms of statistical power. Although the target sample size (n = 63) was not reached, the statistical power achieved with the obtained sample (n = 48) exceeded the accepted threshold.

#### ② Missing Data

No missing values were observed (0.0%) across all nine items of the MPIIQM-J (four pain intensity items and five pain interference items) or across any items of the concurrently administered BPI and QuickDASH. This value was below the acceptable threshold of less than 3% reported in the original publication (Berque et al., 2014).

#### ③ Structural Validity

Prior to conducting factor analysis, the suitability of the data was examined. Bartlett’s test of sphericity yielded χ^2^ (36) = 339.82, p < .001, confirming the presence of a statistically significant correlation structure among the variables. The overall KMO value was .856, substantially exceeding the threshold of .50 recommended by Kaiser (1974) for item retention, with the lowest individual item value of .801 also indicating sufficient adequacy. These results confirmed that the present data were suitable for factor analysis.

Exploratory factor analysis (EFA) was conducted using the principal axis factoring (PAF) method. The number of factors was determined using the Kaiser criterion (eigenvalue ≥ 1), and Varimax rotation was applied. The analysis yielded two factors with eigenvalues exceeding 1 (F1 = 5.27, F2 = 1.64), and a two-factor solution was retained (see Table 1). These two factors accounted for 76.8% of the total variance (F1: 39.1%, F2: 37.6%).

**Table 1.**
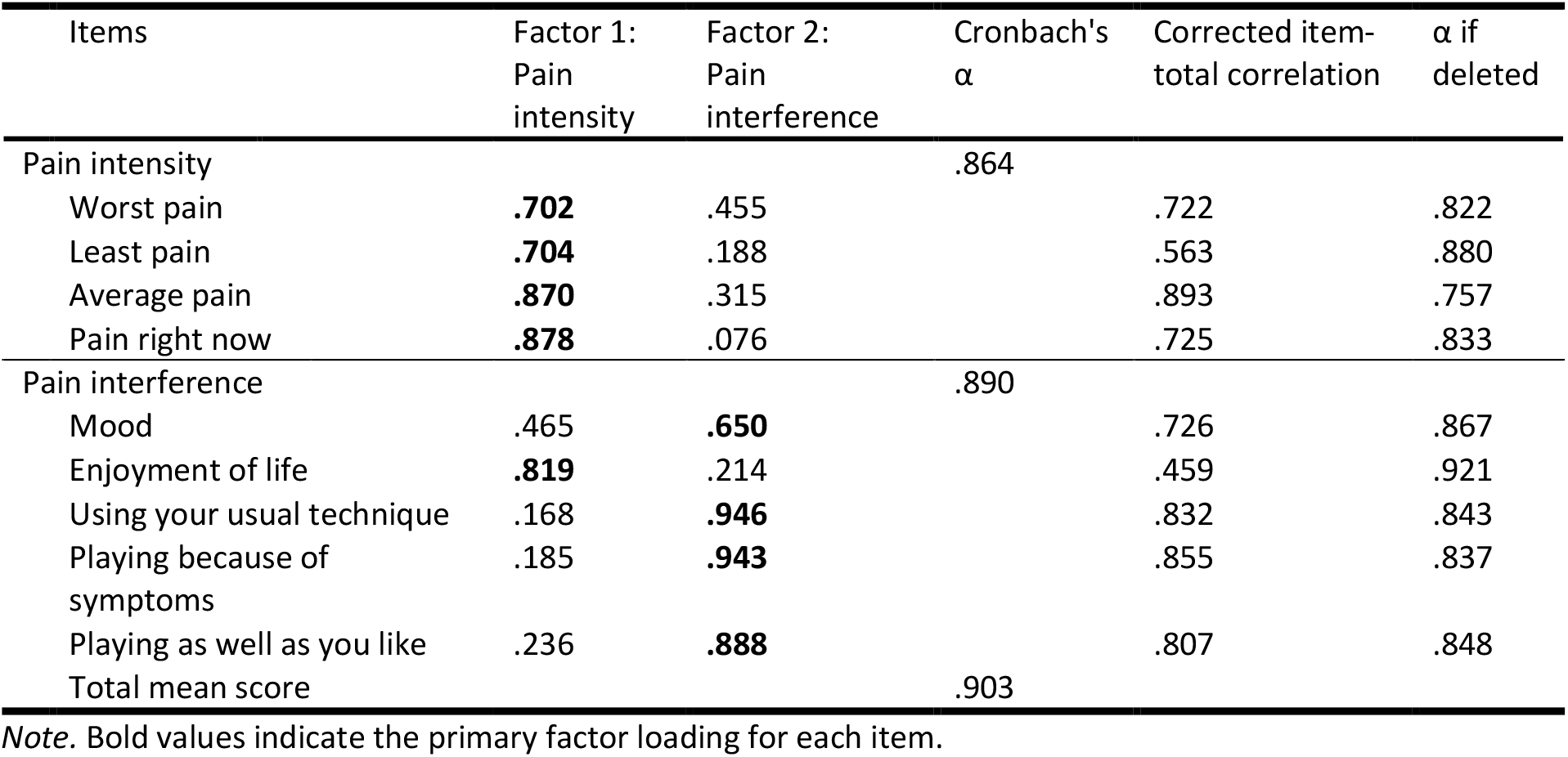
Factor loadings for 9 items of the MPIIQM-J following exploratory factor analysis.

| Items | Factor 1:<br>Pain<br>intensity | Factor 2:<br>Pain<br>interference | Cronbach's<br>$\alpha$ | Corrected item-total correlation | $\alpha$ if<br>deleted |
| --- | --- | --- | --- | --- | --- |
| Pain intensity |  |  | .864 |  |  |
| Worst pain | **.702** | .455 |  | .722 | .822 |
| Least pain | **.704** | .188 |  | .563 | .880 |
| Average pain | **.870** | .315 |  | .893 | .757 |
| Pain right now | **.878** | .076 |  | .725 | .833 |
| Pain interference |  |  | .890 |  |  |
| Mood | .465 | **.650** |  | .726 | .867 |
| Enjoyment of life | **.819** | .214 |  | .459 | .921 |
| Using your usual technique | .168 | **.946** |  | .832 | .843 |
| Playing because of<br>symptoms | .185 | **.943** |  | .855 | .837 |
| Playing as well as you like | .236 | **.888** |  | .807 | .848 |
| Total mean score |  |  | .903 |  |  |

Note. Bold values indicate the primary factor loading for each item.

Notably, the order of the factors was reversed in the Japanese version compared with the original publication and previous adaptations such as the German (MPIIQM-G) and Portuguese (MPIIQM-Pt) versions. Although this reversal has also been reported in the Polish version (MPIIQM-P), it reflects the relative magnitude of variance explained under Varimax rotation and does not compromise the content validity of the factors (Cygańska et al., 2021).

#### ④ Internal Consistency

Internal consistency for each subscale was examined using Cronbach’s α (see Table 1). The α coefficient was .864 for the pain intensity subscale, .890 for the pain interference subscale, and .903 for all nine items combined, confirming high internal consistency across all scales. Corrected item-total correlations ranged from .563 to .893 for the pain intensity subscale and from .459 to .855 for the pain interference subscale. Two items showed an increase in α upon deletion: “Least pain” (α = .880) and “Enjoyment of life” (α = .921). However, neither item had a meaningful adverse effect on the overall α, and the reliability of the scale was considered well maintained.

#### ⑤ Test-Retest Reliability

To assess test-retest reliability, the questionnaire was re-administered in person three to five days after the initial completion, yielding paired data from 30 participants (60 observations in total). However, only 9 of these participants both responded “yes” to Q11 or Q12 of the MPIIQM and had no missing values, rendering calculation of the intraclass correlation coefficient (ICC) impossible.

#### ⑥ Convergent Validity

Correlation coefficients between each subscale of the MPIIQM-J and the BPI, QuickDASH, and QuickDASH Performing Arts Module are presented in Table 2. The MPIIQM-J pain intensity subscale demonstrated a strong and significant correlation with BPI pain severity (r = .71), and moderate significant correlations with BPI pain interference (r = .41), QuickDASH (Q.1–Q.11) (r = .50), and the QuickDASH Performing Arts Module (r = .41). The MPIIQM-J pain interference subscale showed a moderate to strong significant correlation with the QuickDASH Performing Arts Module (r = .62) and a moderate significant correlation with QuickDASH (Q.1– Q.11) (r = .46). BPI pain severity yielded a low to moderate significant correlation (r = .42), whereas BPI pain interference did not reach statistical significance (r = .27). These findings indicate that both subscales of the MPIIQM-J broadly capture the related constructs of pain intensity and interference with performance activities, providing partial support for convergent validity.

**Table 2.** Convergent and divergent validity of the MPIIQM-J subscales with the subscales of the BPI and QuickDASH.

| Items | MPIIQM-J (pain intensity) | MPIIQM-J (pain interference) |
| --- | --- | --- |
| BPI (pain severity) | .71** | .42** |
| BPI (pain interference) | .41** | .27 |
| QuickDASH (Q.1-Q.11) | .50** | .46** |
| QuickDASH (performance module) | .41** | .62** |

Note. \*\* $p$ < .01. All correlations are Pearson's $r$ . $n$ = 48. One correlation (BPI pain interference × pain interference) did not reach statistical significance.

#### ⑦ Comparison between In-person and Online Survey

A comparison of item scores between the in-person survey (n = 23) and the online survey (n = 25) is presented in Table 3. Mean scores were higher in the online survey than in the in-person survey for all items except “Enjoyment of life.” The mean scores for the pain intensity subscale were M = 3.28 (SD = 1.37) for the in-person group and M = 3.63 (SD = 1.78) for the online group; for the pain interference subscale, M = 4.27 (SD = 2.47) and M = 4.59 (SD = 2.32), respectively. Mann-Whitney U tests with Holm correction revealed no statistically significant differences across any of the nine items, and overall differences between the two groups were small (total mean effect size: r = .045).

**Table 3.** Comparison of Mean Item Scores (SD) Between In-Person and Online Surveys.

| Items | M (SD) |  |
| --- | --- | --- |
| | In-person<br>($n = 23$) | Online<br>($n = 25$) |
| Pain intensity | 3.28 (1.37) | 3.63 (1.78) |
| Worst pain | 5.26 (1.94) | 5.76 (1.76) |
| Least pain | 1.22 (1.09) | 1.76 (1.96) |
| Average pain | 3.83 (1.59) | 4.00 (1.89) |
| Pain right now | 2.83 (1.87) | 3.00 (2.65) |
| Pain interference | 4.27 (2.47) | 4.59 (2.32) |
| Mood | 4.13 (3.24) | 4.68 (2.53) |
| Enjoyment of life | 3.30 (2.57) | 3.20 (2.81) |
| Using your usual technique | 4.78 (3.10) | 5.00 (2.50) |
| Playing because of symptoms | 4.61 (3.19) | 4.88 (2.54) |
| Playing as well as you like | 4.52 (3.20) | 5.20 (2.97) |
| Total mean score | 3.83 (1.85) | 4.16 (1.88) |

#### ⑧ Relationship Between Pain and Participant Characteristics

To exploratorily examine trends across instrument groups, Kruskal-Wallis tests were conducted (see Table 4). No statistically significant differences were found among instrument groups (Piano, Strings, Wind, Brass, and Percussion) for age, BMI, MPIIQM-J pain intensity, pain interference, total score, or symptom counts from the Comprehensive Survey of Living Conditions (all p > .05). Descriptively, strings and wind players tended to show slightly higher pain interference scores (M = 5.18 and 5.12, respectively) and higher numbers of symptoms selected from the Comprehensive Survey of Living Conditions (both M = 8.00) compared with other groups, whereas percussion players showed the lowest values across nearly all indices. However, given the small sample sizes for wind (n = 5) and percussion (n = 5) groups, these trends should be interpreted with caution.

**Table 4.** Comparison of Characteristics by Instrument Group.

| Items, M (SD) | Piano<br>($n = 18$) | Strings<br>($n = 12$) | Wind<br>($n = 5$) | Brass<br>($n = 8$) | Percussion<br>($n = 5$) | H | p |
| --- | --- | --- | --- | --- | --- | --- | --- |
| Age | 20.83 (1.38) | 21.33 (1.15) | 21.20 (1.48) | 21.75 (1.75) | 22.40 (3.65) | 2.365 | .669 |
| BMI | 19.78 (1.64) | 20.40 (2.52) | 20.32 (2.31) | 21.60 (1.96) | 20.88 (1.80) | 4.850 | .303 |
| MPIIQM-J Pain intensity | 3.50 (1.28) | 3.58 (2.17) | 3.45 (2.09) | 3.31 (1.25) | 3.30 (1.59) | 0.196 | .996 |
| MPIIQM-J Pain interference | 4.71 (1.96) | 5.18 (2.51) | 5.12 (3.04) | 3.58 (2.22) | 2.36 (2.30) | 5.565 | .234 |
| MPIIQM-J Total | 4.17 (1.50) | 4.47 (2.25) | 4.38 (2.39) | 3.46 (1.50) | 2.78 (1.95) | 3.112 | .539 |
| No. of symptoms reported | 5.33 (3.66) | 8.00 (5.67) | 8.00 (7.28) | 5.50 (2.88) | 4.20 (3.83) | 2.899 | .575 |

*Note.* M = mean; SD = standard deviation. Values are presented as M (SD) unless otherwise indicated. H = Kruskal-Wallis H statistic; p = asymptotic significance (two-tailed). No statistically significant differences were found across instrument groups for any variable (all $p > .05$ ). BMI = body mass index. No. of symptoms reported = Comprehensive Survey of Living Conditions (Ministry of Health, Labour and Welfare, Japan, 2022).

#### ⑨ Descriptive Analysis

To compare the prevalence of pain and symptoms with data from the general Japanese population, results from the Comprehensive Survey of Living Conditions conducted by the Ministry of Health, Labour and Welfare, Japan were utilized. Although this survey is conducted annually, health-related questions are administered once every three years; accordingly, the most recent available data from 2022 were used in the present analysis. As the survey data are aggregated in five-year age bands, the 20–24 age group was selected as the reference, corresponding to the mean age of the present sample (M = 21.31, SD = 1.74). The survey comprises 41 items asking respondents to select any subjective symptoms of concern, with multiple responses permitted. Binomial tests were conducted to compare the prevalence of each symptom among students in HMEIs (n = 226) against the national population proportions as the reference, with Holm correction applied for multiple comparisons. It should be noted that the full sample (n = 226) was used for this comparison rather than the symptomatic subgroup, in order to ensure representativeness of the population when comparing symptom prevalence with national data.

The results of the comparison of symptom prevalence between the Comprehensive Survey of Living Conditions and students in HMEIs are presented in Tables 5 and 6.

**Table 5.**
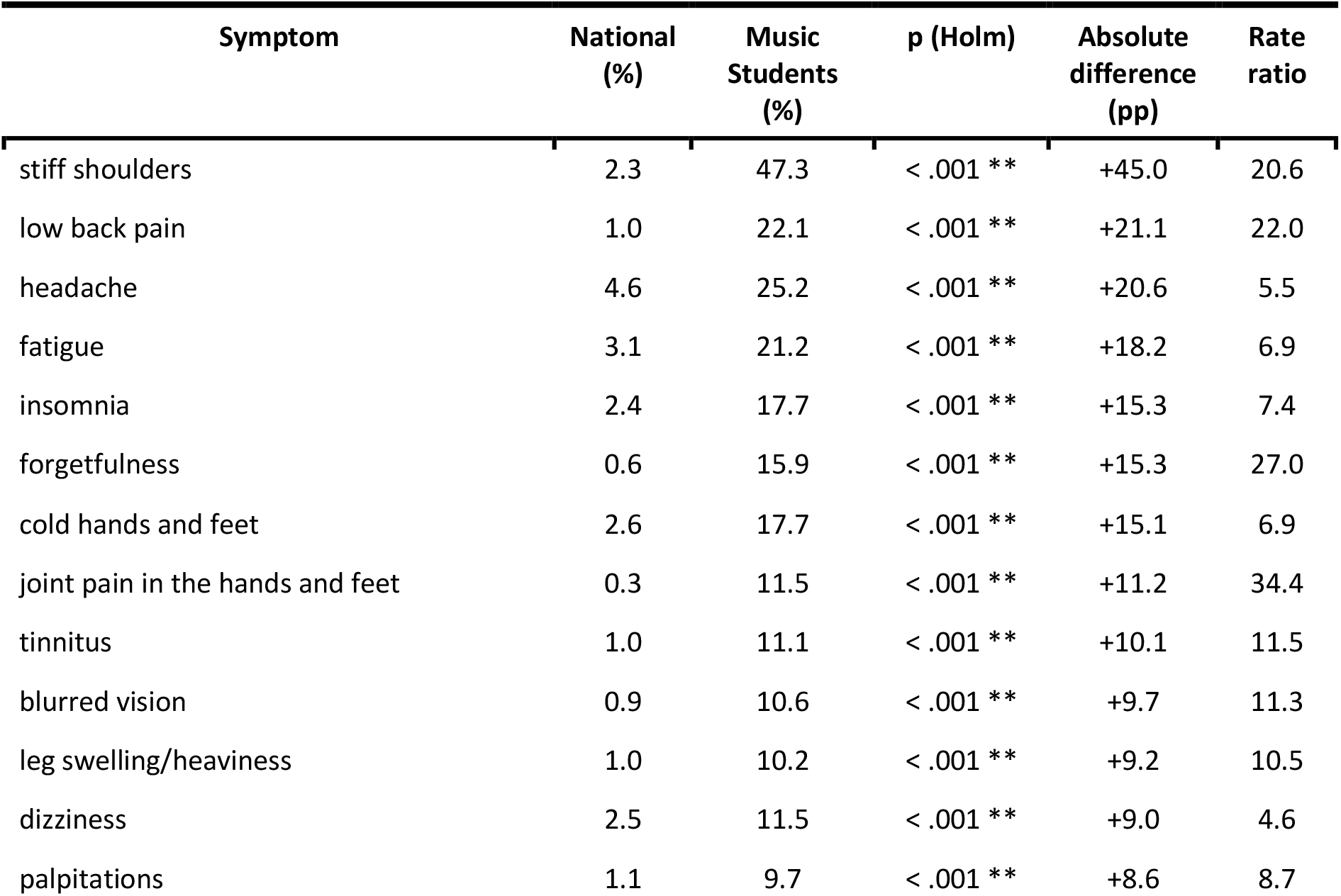
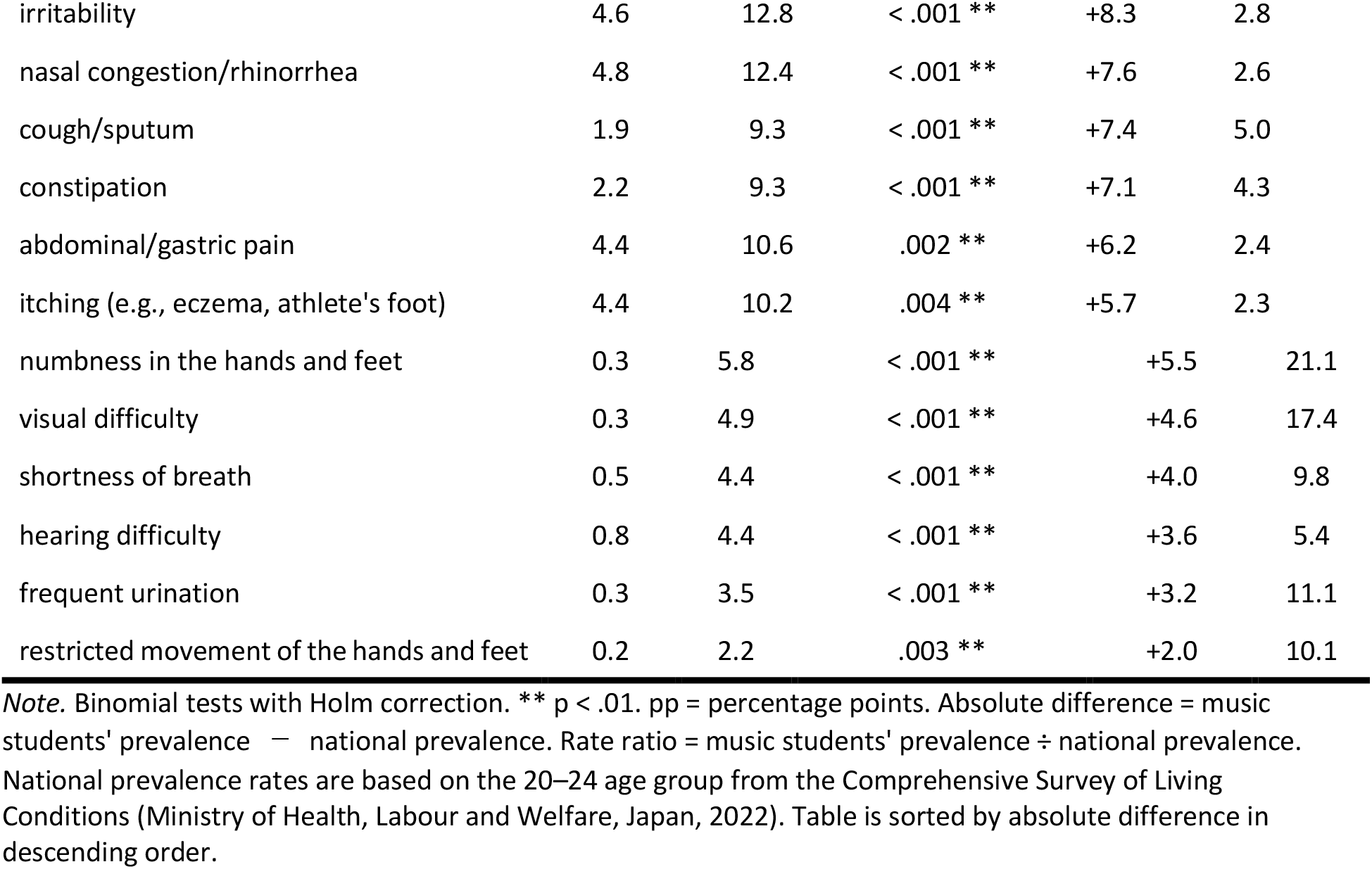
Comparison of Symptom Prevalence Between the Comprehensive Survey of Living Conditions (Ages 20–24) and students in Higher Music Education Institutions (n = 226, M = 20.78, SD = 2.84); Symptoms with Significant Differences (25 Items)

**Table 6.** Comparison of Symptom Prevalence Between the Comprehensive Survey of Living Conditions (Ages 20–24) and Students in Higher Music Education Institutions (n = 226, M = 20.78, SD = 2.84); Symptoms with No Significant Differences (16 Items)

| Symptom | National (%) | Music Students (%) |
| --- | --- | --- |
| Menstrual irregularity/dysmenorrhea | 12.5 | 11.1 |
| Diarrhea | 5.0 | 8.0 |
| Stomach heaviness/heartburn | 2.3 | 4.9 |
| Loss of appetite | 2.0 | 4.4 |
| Toothache | 3.6 | 4.4 |
| Skin rash (e.g., urticaria, eruptions) | 6.1 | 4.0 |
| Injuries (e.g., cuts, burns) | 4.2 | 3.1 |
| Wheezing | 1.1 | 2.2 |
| Swollen or bleeding gums | 0.6 | 1.8 |
| Fever | 3.4 | 1.8 |
| Chest pain | 2.2 | 1.8 |
| Hemorrhoids (pain, bleeding, etc.) | 2.2 | 0.9 |
| Difficulty urinating/painful urination | 0.6 | 0.0 |
| Urinary incontinence | 0.8 | 0.0 |
| Difficulty chewing | 1.4 | 0.4 |
| Fractures/sprains/dislocations | 1.7 | 0.4 |

*Note.* No statistically significant differences were found (binomial tests with Holm correction, all $p > .05$ ). National prevalence rates are based on the 20–24 age group from the Comprehensive Survey of Living Conditions (Ministry of Health, Labour and Welfare, Japan, 2022). Table is sorted by Music Students in descending order.

Binomial tests with Holm correction revealed that students in HMEIs showed significantly higher symptom prevalence than the general population of the same age group in 25 of the 41 items. Symptoms with significant differences were observed across the following categories: Musculoskeletal & Peripheral Neurological (stiff shoulders, low back pain, joint pain in the hands and feet, numbness in the hands and feet, restricted movement of the hands and feet), Psychological, Neurological & General Symptoms (fatigue, forgetfulness, insomnia, irritability, headache, dizziness), Cardiorespiratory-related Symptoms (cold hands and feet, leg swelling/heaviness, palpitations, shortness of breath), Visual & Auditory Symptoms (tinnitus, blurred vision, visual difficulty, hearing difficulty), Upper Airway & Respiratory Symptoms (nasal congestion/rhinorrhea, cough/sputum), and Gastrointestinal, Urinary & Dermatological Symptoms (constipation, frequent urination, abdominal/gastric pain, itching).

The symptoms with the largest absolute differences were stiff shoulders (45.0 percentage points), low back pain (21.1 percentage points), and headache (20.6 percentage points). The highest prevalence ratios were observed for joint pain in the hands and feet (34.4-fold), forgetfulness (27.0-fold), low back pain (22.0-fold), stiff shoulders (20.6-fold), and numbness in the hands and feet (21.1-fold). In contrast, no symptoms were identified for which the prevalence among students in HMEIs was significantly lower than that of the general population.

## 4 Discussion

### 4.1. Factor Structure of the MPIIQM-J and Comparison with Previous Studies

The results of this study indicate that the MPIIQM-J exhibits a two-factor structure (Pain intensity and Pain interference) consistent with the original version (Berque et al., 2014) and other language versions, explaining 76.8% of the total variance. Although the order of the factors differed from that reported in the original, German, and Portuguese versions, this difference may be attributable to variations in the proportion of explained variance under Varimax rotation. A similar reversal has also been reported in the Polish version (Cygańska et al., 2021). Importantly, the content validity of the factors appeared to be preserved, and no substantial issues in interpretation were identified. With regard to the study population, the present study focused on students in HMEIs, which differs from previous studies that primarily targeted professional orchestral musicians. Nevertheless, the observed factor structure and overall trends in psychometric indices appeared to be broadly consistent with those reported in prior research. According to communication with the original authors, the MPIIQM is not necessarily restricted to professional orchestral musicians and may be applicable to a broader population, including amateur musicians and students in HMEIs. The findings of the present study are in line with this perspective and may suggest potential applicability of the MPIIQM-J beyond the originally intended population. However, given the differences in sample characteristics, caution is warranted when considering the generalizability of these findings. While the results may provide useful insights into playing-related health problems, further validation in diverse musician populations is required.

### 4.2. Relatively Low Prevalence of Playing-Related Symptoms: Cultural and Cognitive Factors

In the face-to-face survey (n = 200), only 27 participants (13.5%) reported symptoms related to playing before exclusion. This relatively low prevalence differs substantially from the rate of 50% or higher identified in the literature review conducted by Akaike (2026), and the reasons for this discrepancy warrant careful consideration.

First, suppression of pain expression in Asian cultural contexts may have contributed to this finding. Liao et al. (2016) reported that many Asian participants did not express pain to others and, when they did, often experienced regret or shame, conceptualizing this tendency as “culturally imposed stoicism.” In addition, within the musician-specific context, it has been suggested that disclosing playing-related pain may entail a risk of losing professional opportunities, and that a strong orientation toward excellence may further discourage self-disclosure of difficulties (Ackermann, 2017; Détári et al., 2026).

Second, issues related to the interpretation of the concept of “interference with performance” were suggested. In the preliminary interviews conducted after the face-to-face survey, a shared cognitive framework emerged among respondents who reported pain on the BPI but did not endorse symptoms on the MPIIQM. Specifically, these individuals appeared to interpret their condition as “I have pain, but I can continue performing; therefore, it does not interfere with my performance.” Although the MPIIQM defines playing-related musculoskeletal problems as symptoms that interfere with the usual level of performance, this criterion may have been interpreted as equivalent to a complete inability to perform. These findings suggest that, when using the MPIIQM-J, it is essential to incorporate a clear explanation and confirmation process regarding the definition of “playing-related musculoskeletal problems” to ensure consistent interpretation among respondents.

Against this background, the required sample size for factor analysis could not be obtained from the face-to-face survey. Therefore, in the second online survey, additional data were collected by restricting participants to those who had experienced pain during performance within the past month. As a result, the mean scores in the online survey exceeded those of the face-to-face survey across all items (see Table 3). This pattern may be explained by the fact that participants in the online survey were individuals who recognized both the presence of pain during performance and its associated interference.

### 4.3. Health Risks Students in Higher Music Education Institutions: Comparison with the National Survey

When the physical and psychological symptoms reported by students in HMEIs were compared with those of the general population of the same age based on the Comprehensive Survey of Living Conditions marked differences were observed. Of the 41 items, 25 showed significantly higher prevalence rates among students in HMEIs, with particularly pronounced differences in musculoskeletal-related symptoms (stiff shoulders, low back pain, joint pain in the hands and feet, numbness in the hands and feet, and restricted movement). These symptoms are directly related to performance-related physical demands and suggest the accumulation of physical load due to prolonged practice and repetitive movements. In addition, symptoms categorized as Psychological, Neurological & General Symptoms (e.g., fatigue, insomnia, and forgetfulness), as well as auditory-related symptoms such as tinnitus and hearing difficulties, were also significantly more prevalent among students in HMEIs. These findings underscore the substantial health risks faced by this population.

Taken together, the results highlight the need for health management and preventive educational interventions within music education institutions, and suggest that the MPIIQM-J may serve as a useful screening tool in such contexts.

### 4.4. Need for an Additional Explanatory Statement on the Interpretation of the Definition of Playing-Related Pain

By comparing responses using identical questionnaire items from the Comprehensive Survey of Living Conditions, it was demonstrated that students in HMEIs exhibit a markedly higher prevalence of musculoskeletal-related symptoms than the general population of the same age group. However, this finding appears inconsistent with the fact that 173 cases (86.5%) from the initial face-to-face survey were excluded from the analysis. To explore this discrepancy, the number of participants reporting pain or problems in the hands and lower back was examined using both the MPIIQM-J and the BPI. Among the analyzed sample, 13 participants (27.8%; hands: 10, lower back: 3) reported such symptoms on the MPIIQM-J, while 9 participants (18.7%; hands: 7, lower back: 2) did so on the BPI. In contrast, among the 152 participants who were ultimately excluded, 5 (3.2%; hands: 3, lower back: 2) reported symptoms on the MPIIQM-J, and 19 (12.5%; hands: 10, lower back: 9) reported symptoms on the BPI. These results indicate that a certain proportion of excluded participants also experienced musculoskeletal pain related to performance.

As noted above, qualitative findings suggest the presence of a cognitive framework in which individuals may experience pain but do not endorse it as playing-related musculoskeletal pain. The comparison using identical questionnaire items from the Comprehensive Survey of Living Conditions provides quantitative support for this interpretation. Accordingly, these findings highlight the need to include an additional explanatory statement for the definition of playing-related musculoskeletal problems presented immediately before Q9 of the MPIIQM-J, in order to minimize variability in interpretation among respondents. The MPIIQM-J is available from the corresponding author upon request.

### 4.5. Internal Consistency : Positioning of “Least pain” and “Enjoyment of life”

Cronbach’s α coefficients for each subscale were .864 for Pain intensity, .890 for Pain interference, and .903 for all nine items combined, indicating high internal consistency across the scale. However, for the two items “Least pain” and “Enjoyment of life,” an increase in α was observed when each item was deleted. For “Least pain,” compared with the other three items (worst, average, and current pain), the score may diverge when participants exhibit relatively small fluctuations in pain, resulting in weaker correlations within the subscale. Regarding “Enjoyment of life,” whereas the other Pain interference items (performance technique, difficulty in performance, and ability to perform as intended) assess specific interference with performance activities, this item captures the impact on overall life enjoyment. As such, its conceptual linkage to the performance-specific context may be relatively weaker. Importantly, these tendencies are not unique to the MPIIQM-J. An increase in α upon item deletion for “Least pain” has also been reported in the German (MPIIQM-G), Brazilian Portuguese (MPIIQM-Br), and European Portuguese (MPIIQM-Pt) versions, while similar findings for “Enjoyment of life” have been reported in the Brazilian Portuguese (MPIIQM-Br) and European Portuguese (MPIIQM-Pt) versions. Therefore, these patterns should be understood as structural characteristics of the MPIIQM. Given that neither item had a substantial negative impact on α and that overall reliability remained sufficient, the removal of these items was not considered justified.

### 4.6. Convergent Validity

Convergent validity was supported. A significant correlation was observed between the MPIIQM-J pain intensity subscale and the BPI pain severity scale (r = .71), meeting the a priori hypothesis (r > 0.70). In addition, the MPIIQM-J pain interference subscale showed a correlation of r = .62 with the QuickDASH Performing Arts Module, which fell within the expected range of the a priori hypothesis (r = 0.50–0.70).

### 4.7. Limitations and Future Directions

In addition to the limitations described above, several issues warrant further consideration.

First, test–retest reliability (intraclass correlation coefficient [ICC]) could not be calculated, as only nine of the 30 retest respondents met the criteria for analysis (i.e., complete data and affirmative responses to MPIIQM Q11 or Q12). While a similar limitation was reported in the MPIIQM-P (Cygańska et al., 2021), the present findings suggest that, in the case of the MPIIQM-J, cultural and cognitive factors may have contributed to an underestimation of symptoms, thereby reducing the number of respondents identified as symptomatic. Future studies should address this issue by recruiting participants with confirmed playing-related problems.

Second, although statistical power was adequate, the stability of the factor structure was not formally assessed and should be confirmed in future studies with larger samples.

Third, in the exploratory factor analysis, the item “Enjoyment of life” showed a high factor loading (.819) on the Pain intensity factor. This may reflect differences in how respondents interpret the relationship between emotional distress and pain-related constructs. This issue remains to be examined in further validation studies of the MPIIQM-J.

### 4.8. Implications for Measurement of Playing-Related Musculoskeletal Disorders

Despite these methodological limitations, the present study provides important insights into the assessment of playing-related musculoskeletal disorders. In particular, the discrepancy observed between responses to the BPI and the MPIIQM-J suggests that the identification of symptoms may depend not only on the presence of pain but also on how individuals interpret its impact on performance within a performance-specific context. This finding highlights the potential influence of cognitive and contextual factors on self-reported outcomes and suggests that the assessment of PRMDs may be sensitive to differences in how respondents conceptualize performance-related interference. From this perspective, the present results may contribute to a more nuanced understanding of measurement issues in musician-specific health research.

## 5. Conclusions

The MPIIQM-J was developed through a translation process that incorporated linguistic validation by professional musicians and subsequent evaluation of validity by students in HMEIs. The MPIIQM-J reproduced the two-factor structure reported in previous studies and demonstrated adequate internal consistency and convergent validity. However, test–retest reliability (intraclass correlation coefficient [ICC]) could not be calculated, as only 9 of the 30 retest respondents met the criteria for analysis. In addition, in the exploratory factor analysis, the item “Enjoyment of life” showed a high loading on the Pain intensity factor, which remains an issue to be examined in future validation studies. Furthermore, the findings suggested that the definition of playing-related musculoskeletal problems may be interpreted inconsistently among respondents. Therefore, it is recommended that a clear explanatory statement of the definition be provided when administering the MPIIQM-J. Accordingly, an explanatory statement developed by the authors of the MPIIQM-J is included as an Appendix. Taken together, the scale is expected to be useful in educational and clinical settings for both amateur and professional instrumental musicians. Nevertheless, further psychometric evaluation, including verification of test–retest reliability, is warranted.

## Data Availability

The data in this study are potentially sensitive health information. We do not have ethics approval to share them outside of the research team.

